# Systematic discovery of gene-environment interactions underlying the human plasma proteome in UK Biobank

**DOI:** 10.1101/2023.10.26.23297604

**Authors:** Robert F. Hillary, Danni A. Gadd, Zhana Kuncheva, Tasos Mangelis, Tinchi Lin, Kyle Ferber, Helen McLaughlin, Heiko Runz, Biogen Biobank Team, Riccardo E. Marioni, Christopher N. Foley, Benjamin B. Sun

## Abstract

Understanding how gene-environment interactions (GEIs) influence the circulating proteome could aid in biomarker discovery and validation. The presence of GEIs can be inferred from single nucleotide polymorphisms that associate with phenotypic variability - termed variance quantitative trait loci (vQTLs). Here, vQTL association studies are performed on plasma levels of 1,468 proteins in 53,752 UK Biobank participants. A set of 683 independent vQTLs are identified across 571 proteins, all of which are newly discovered. They include 65 variants that lack conventional additive main effects on protein levels. Over 1,400 GEIs are identified between 142 proteins and 101 lifestyle and metabolic exposures. GEI analyses uncover biological mechanisms that explain why 13/65 vQTL-only sites lack corresponding main effects. Stratified analyses also highlight how age, sex and genotype can interact to modify relationships between biomarkers and health-related traits. This study establishes the most comprehensive database yet of vQTLs and GEIs for the human proteome.

## Introduction

High-throughput proteomic analyses enable scalable biomarker discovery for complex disease states ^1^. A growing number of studies have catalogued genetic influences on the human plasma proteome ^2–6^. Sequence variants associated with protein abundances are termed protein quantitative trait loci (or pQTLs) and their colocalisation with disease-associated variants guides the identification of pathogenic molecular pathways, aiding drug and biomarker validation ^7–9^. However, the influences of environmental factors and, in particular, gene-environment interactions (or GEIs) on the human plasma proteome have remained understudied. Determining whether environmental exposures modify genetic associations with protein abundances should provide additional, nuanced insights into protein biology and personalised medicine strategies.

GEIs most commonly arise when genotype groups at a locus show differential associations between an environmental exposure and a phenotype of interest (e.g. protein levels) ^10,11^. There has been relatively limited success in identifying GEIs due to their small effect sizes and challenges in accurately recording multiple environmental exposures over the life course ^12,13^. Using all genome-wide genetic variants and hundreds of potential environmental modifiers to test for GEIs also imposes a significant multiple testing burden.

A GEI can manifest in the form of differences in the variance of a given trait across genotypes at a locus (**Fig. 1**). Therefore, one strategy to infer the presence of a GEI is to perform genome-wide scans for these loci, which are defined as variance quantitative trait loci or vQTLs ^14,15^. This is in contrast with pQTLs, which associate with differences in mean protein levels across genotype groups. Of note, variants that associate with mean differences in traits (i.e. pQTLs) have been referred to as additive main effect or simply, main effect loci in the vQTL literature ^16,17^. Studies have identified vQTLs for lifestyle and cardiopulmonary traits such as blood pressure and body mass index ^15,16,18,19^. These studies have also shown that the power to detect GEIs is enhanced when restricting the genetic search space to vQTLs instead of all genome-wide variants or to QTLs with additive main effects on the outcome (analogous to pQTLs) ^15–17^. Westerman *et al.* (2022) recently applied the two-stage approach of vQTL discovery and GEI testing to serum cardiometabolic biomarkers, which included 10 proteins. However, the vQTL architecture for most human proteins remains undescribed thereby hindering systematic screens for GEIs that may impact their circulating levels. Identifying vQTLs and associated GEIs in this context could further guide predictions on the safety and efficacy of protein biomarkers and drug targets.

**Fig. 1.**
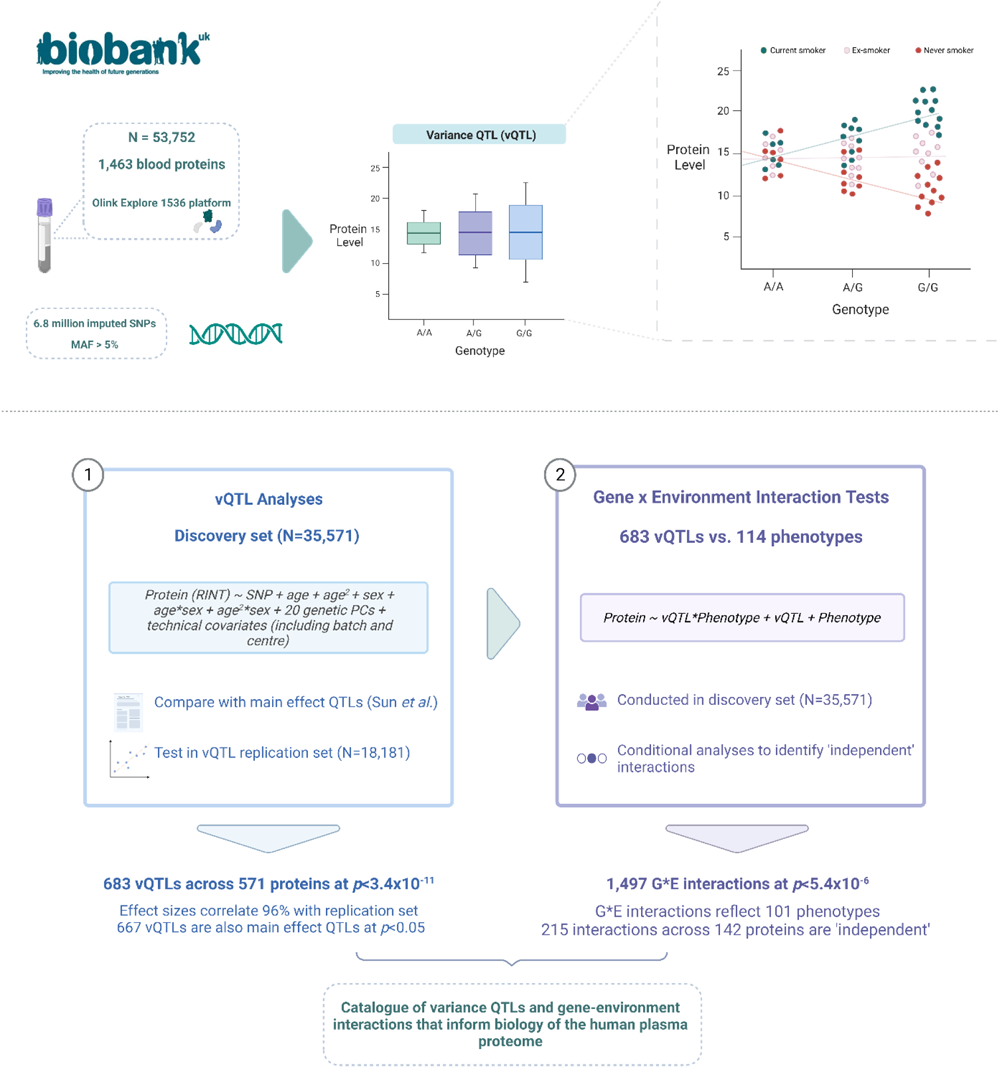
Overview of study design for variance QTL analyses in The UK Biobank Pharma Proteomics Project sample. Top panel: Plasma levels of 1,463 proteins (measured by 1,472 Olink analytes) and genotype data were available for 53,752 participants in UK Biobank following quality control. Variance QTL (vQTL) analyses were performed using 6.8 million imputed SNPs to detect loci that associated with differential variances in protein levels across genotypes. Gene-environment interactions (GEIs) can manifest as differences in the variance of a trait (e.g. protein levels) across genotypes at a given polymorphism. In this example, which uses fictitious data, the G-allele positively correlates with protein levels in one sub-group of the sample (current smokers, shown in teal). A negative correlation is observed in never smokers (shown in red). There is no correlation in ex-smokers (shown in peach). Therefore, genotype at this locus interacts with the exposure (e.g. smoking status), which creates a mean-based interaction effect. The effect underlies the dispersion of the data in G-allele carriers and in turn gives the appearance of a vQTL. **Bottom panel:** The independent discovery and replication sets consisted of 35,571 and 18,181 participants, respectively. Effect sizes and *p*-values for variance QTLs were compared against those from a recent main effect QTL analysis on the same proteins and sample by Sun *et al*. The use of vQTLs for GEI tests can greatly reduce computational burden. Therefore, we tested whether protein levels were associated with an interaction between their vQTLs and a broad range of health-related phenotypes in UK Biobank. A large number of phenotypes were correlated with one another (e.g. adiposity-related traits). Stepwise conditional analyses were employed in order to identify ‘independent’ interactions. MAF, minor allele frequency; GEI, gene-environment interactions; vQTL, variance quantitative trait locus. Image created using Biorender.com.

In this study, we first conduct genome-wide vQTL association studies on plasma levels of 1,463 Olink proteins in up-to-53,752 UK Biobank participants. GEI associations are then comprehensively screened using vQTL loci identified in stage one and over 100 lifestyle and metabolic exposures (see **Fig. 1** for a summary of the study design). vQTL variants are cross-referenced with a recent pQTL (or main effect QTL) study using the same sample, highlighting novel aspects of protein biology that would not have otherwise been captured by conventional additive GWAS models ^4^. GEI association tests are also repeated using pQTLs to assess whether there is an enrichment in GEI discovery when restricting the genetic search space to variance effect versus main effect QTLs. Additionally, age-and sex-specific GEI association tests are performed to further inform which strata of the population are most susceptible to a given GEI. This study establishes and prioritises a comprehensive catalogue of variance effects for the human proteome, which others may utilise to investigate GEIs of interest. Further, we detail several, disparate examples that provide biological insights into the complex interplay between the genome, proteome and environment.

## Results

### Discovery of variance QTLs underlying the plasma proteome

In the first stage of the study, Levene’s test was used to perform genome-wide vQTL analyses on blood levels of 1,463 unique proteins. The 1,463 proteins were measured by 1,472 analytes. A Bonferroni-corrected significance threshold was set at *p*<3.4×10^-11^, which reflected the adjustment of *p*<5×10^-8^ (a commonly used threshold in GWAS) ^20^ for 1,463 proteins. There were 274,272 significant vQTL associations across 572 analytes at *p*<3.4×10^-11^. The associations implicated 571 unique proteins as two analytes targeted the same protein. There was limited evidence for genomic inflation (range of λ=[0.9, 1.1], **Supplementary Table 1**). Six hundred and eighty-three independent vQTLs were identified through linkage disequilibrium (LD) clumping (see **Methods**, **Supplementary Table 2**).

Four hundred and seventy-three (70.1%) of the 683 independent vQTLs were *cis* effects (within 1Mb from the gene encoding the protein) and the remaining 210 represented *trans* effects (**Fig. 2A**). The majority of proteins had one independent vQTL (489, 85.6%) and the maximum number of vQTLs per protein was 6 (for FOLR3, **Fig. 2B**). There was an inverse relationship between the logarithms of effect sizes and minor allele frequency (MAF) for both *cis* and *trans* loci (*r*=-0.36 and -0.49, respectively, MAF≥5%, **Fig. 2C**). Six variants were not available for replication testing as they had MAF<5% in the replication set. Of the 677 variants available for testing, 645 (95.3%) were nominally significant (*p*<0.05) and directionally concordant with estimates from the discovery set. Effect sizes were highly correlated between both sets (*r*=0.96, 95% CI=[0.95, 0.97], **Fig. 2D**, **Supplementary Table 3**).

**Fig. 2.**
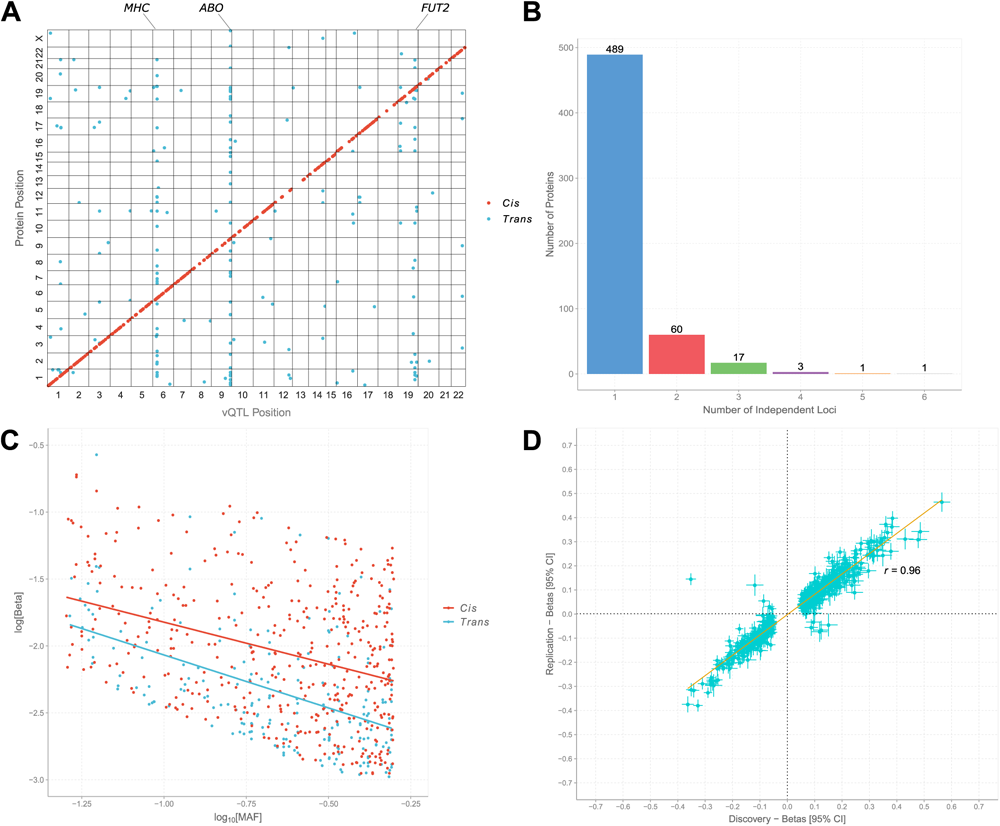
Genome-wide association studies to identify variance QTLs for 1,472 blood protein measures in UK Biobank. **(A)** The 1,472 analytes or measures represented 1,463 unique proteins. A Bonferroni-corrected significance threshold of *p*<3.4×10^-11^ was set. The x-axis represents the chromosomal location of independent *cis* and *trans* vQTLs. The y-axis represents the position of the gene encoding the associated protein. *Cis* (red circles); *trans* (blue circles). **(B)** The number of independent vQTLs per protein. **(C)** Association between the common logarithm of minor allele frequencies and the natural logarithm of absolute effect sizes for *cis* and *trans* vQTLs. *Cis* (red circles and line); *trans* (blue circles and line). **(D)** The discovery and replication sets consisted of 35,571 and 18,181 participants, respectively. Correlations between effect sizes in the discovery and replication sets are shown for vQTLs that were significant in the discovery set. CI, confidence interval; MAF, minor allele frequency; vQTL, variance quantitative trait locus.

### Variance QTLs largely overlap with main effect QTLs for the blood proteome

The majority of vQTLs (618, 90.5%) had main effect *p*-values <3.4×10^-11^ (**Supplementary Table 2**). Only sixteen vQTLs lacked a main effect finding at *p*<0.05. Their *p*-values ranged from 0.07 to 0.98 and five loci were located in the *MHC* region. **Fig. 3A** and **3B** demonstrate examples of vQTL loci with and without marginal main effects, respectively. A reverse look-up strategy showed that only 3.4% of main effect QTLs (299 of 8,857 variants at MAF≥5%) had vQTL *p*-values <3.4×10^-11^ (**Supplementary Table 4**).

**Fig. 3.**
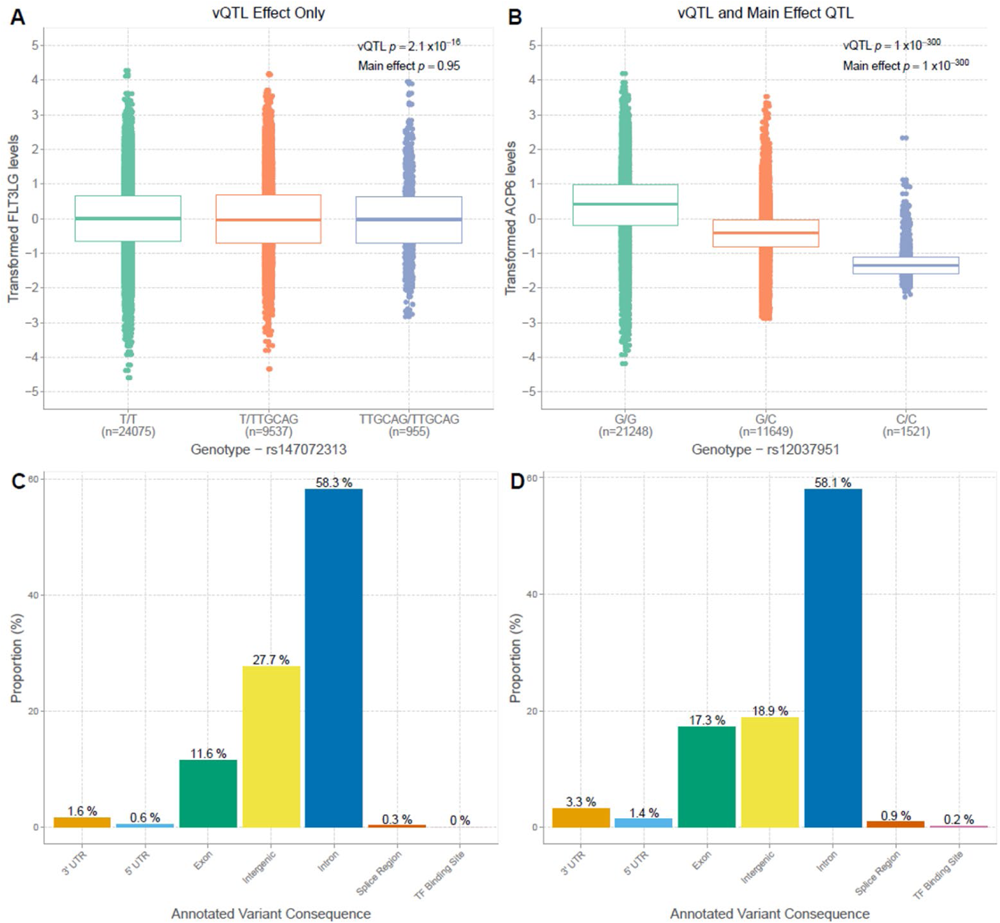
Genetic architectures of main effect and variance QTLs for plasma protein levels. **(A)** Example of variance QTL (rs147072313) without corresponding main effect on protein levels (FLT3LG). **(B)** Example of variance QTL (rs12037951) with significant main effect on protein levels (ACP6). **(C)** Distributions of predicted functional annotation classes for all variance QTLs. Bar height represents the mean proportion of variants within each class. **(D)** Distributions of predicted functional annotation classes for main effect QTLs. TF, transcription factor; UTR, untranslated region; vQTL, variance quantitative trait locus.

**Fig. 3C** and **3D** show the predicted functional classes of vQTLs and main effect QTLs. Most predicted classes exhibited little variation between these QTL types given their substantial overlap. However, a higher proportion of main effect QTLs were annotated to exons than vQTLs (17.3% vs. 11.6%). In balance, a lower proportion of main effect QTLs were annotated to intergenic sites when compared to vQTLs (18.9% vs. 27.7%).

Of note, main effect QTLs and variance QTLs had similar profiles in terms of imputation accuracy and the median INFO scores were 99.3% (interquartile range (IQR)=1.8%) and 99.4% (IQR=1.5%), respectively. The higher proportion of exon annotations for main effect QTLs may reflect variants that affect amino acid sequences and, in turn, antibody-antigen recognition. Indeed, 23% of *cis* main effect associations were protein-altering variants or were in linkage disequilibrium (*r*^2^>0.8) with such variants ^4^. These variants may precipitate differential binding rather than differences in circulating abundances. The effects are likely to influence mean protein measurements across genotypes (i.e. additive main effect or pQTLs) as opposed to differences in the variance, and may contribute to the observed differences in predicted functional classes between main effect and vQTLs.

### Variance QTLs unveil gene-environment interactions dispersed across the plasma proteome

In the second stage of the study, we tested whether protein levels were associated with an interaction between their vQTL(s) and 114 disparate exposures. There were 1,497 GEIs at a Bonferroni-adjusted threshold of *p*<5.4×10^-6^ when vQTLs were used as index variants (*p*<0.05 adjusted for 683 variants and 13.5 effective exposures). Significant GEIs comprised 22.2% of the vQTLs tested (152 variants) and reflected 142 unique proteins (**Supplementary Tables 5 and 6**). The main effect QTL strategy returned 4,036 GEIs at the same threshold. However, these associations encompassed only 4.1% of main effect QTLs (359 of 8,857 variants, **Supplementary Table 7**). Prioritising vQTLs therefore provided a 5.4-fold enrichment in GEI discovery over conventional QTLs at the variant level, and this estimate is in line with previous findings ^15,17^.

Of note, applying the same multiple testing threshold is more liberal for main effect QTLs given their larger number of input variants (683 vs. 8,857) ^17^. Only 2,997 of 4,036 GEIs (73.7%) and 280 sites (3.2%) remained at a tailored threshold of *p*<4.2×10^-7^ (*p*<0.05 adjusted for 8,857 variants and 13.5 effective exposures).

Stepwise conditional analyses suggested that 215 of the 1,497 GEIs with vQTLs were ‘independent’ when further accounting for correlations between phenotypes (see **Methods**, **Fig. 4A**, **Supplementary Table 8**). Eighteen proteins exhibited three or more conditionally significant associations with a maximum of 8 GEIs for PAEP (progestagen-associated endometrial protein or glycodelin, **Fig. 4B**). Conditional GEIs included 62 unique phenotypes. In total, 172/215 GEIs were nominally significant (*p*<0.05) and directionally concordant with estimates from the replication set (**Supplementary Table 9**). Sensitivity analyses for the preparation of phenotypes in vQTL and GEI association tests are detailed in the **Supplementary Note**.

**Fig. 4.**
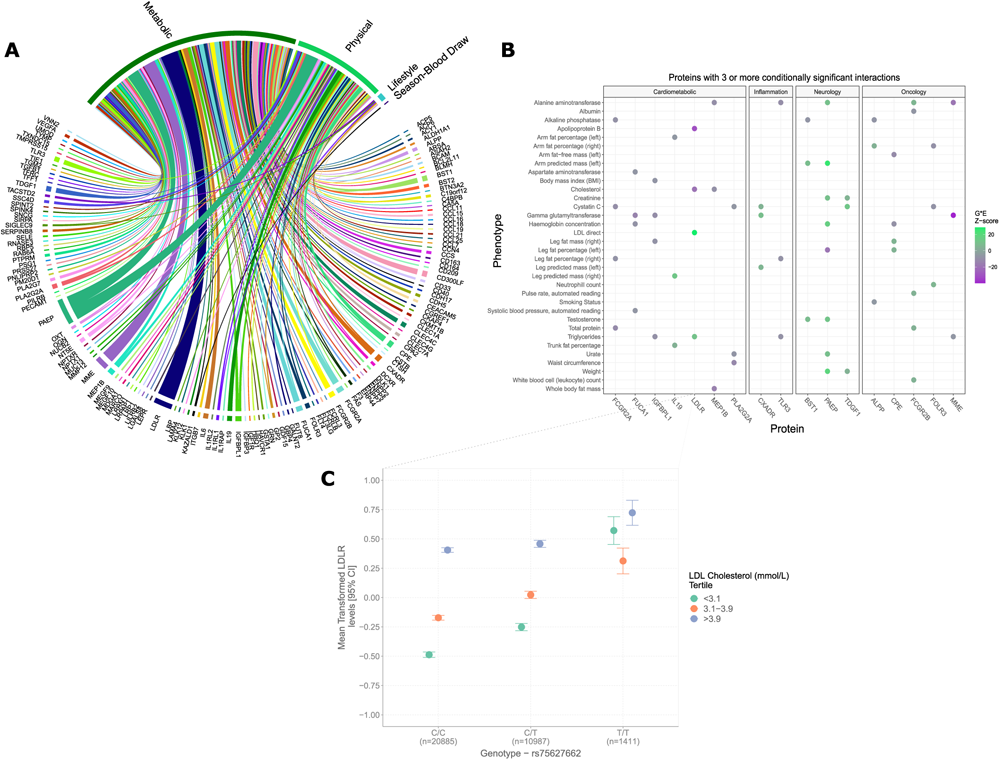
Gene-environment interactions underlying the plasma proteome are pervasive. **(A)** Circos plot showing all conditionally significant GEIs. Only associations that surpassed a Bonferroni-corrected threshold of *p*<5.4×10^-6^ are displayed. Exposure variables are broadly categorised into similar classes for clarity. **(B)** Interaction Z-scores are highlighted only for proteins with three or more conditionally significant GEI effects. Positive Z-scores are shown in green and negative Z-scores are shown in purple. **(C)** Mean transformed LDL receptor or LDLR levels (closed circles) with 95% confidence intervals (vertical bars) when stratified by vQTL genotype (rs75627662) and tertiles of measured blood LDL cholesterol. GEI, gene-environment interaction; LDL, low-density lipoprotein; vQTL, variance quantitative trait locus.

GEIs revealed interactions consistent with known biology, such as associations between LDL receptor (low-density lipoprotein, LDLR) and LDL cholesterol. Our data illustrate genotype-dependent effects for these relationships. For instance, the *trans* variant rs75627662-T located near *APOE* had an additive effect on transformed LDLR levels and served as a vQTL (**Fig. 4C**). There was no correlation between LDLR and cholesterol in individuals homozygotes for the minor T-allele (*r*_minor_=0.005, Pearson’s correlation). However, heterozygotes showed a positive correlation (*r_het_*=0.32) and a stronger association was observed in those homozygous for major (C)-allele (*r*_major_*=*0.41). This underscored a clear GEI that was uncovered by the two-stage strategy.

### Gene-environment interactions are influenced by age and sex differences

It is unclear whether a given interaction remains consistent across an entire study population (i.e. across age ranges and sexes). We therefore sought to highlight GEIs that may be susceptible to age and/or sex differences. Linear regression models were used to test whether protein levels were associated with (i) genotype-by-sex and (ii) genotype-by-age interaction terms. Only the 142 proteins that participated in GEIs were considered. Two proteins (GP2 and TACSTD2) showed age-biased QTLs at *p*<5.4×10^-6^ (matching the threshold used in GEI tests, **Supplementary Table 10**). Twenty-nine proteins associated with a genotype-by-sex interaction at this threshold and the strongest association was observed for PAEP (*p*=4.7×10^-223^).

The association between PAEP and body weight clearly illustrated how GEIs can be accounted for by sex differences. The *cis* rs697449-T variant was a vQTL for PAEP levels and also showed an additive main effect on PAEP levels (**Fig. 5A**). Moderate correlations between PAEP and body weight were observed in T-allele carriers when males and females were analysed together (*r*=0.22 for heterozygotes and T-homozygotes, **Fig. 5A**). However, a weak negative association was observed in major (G)-allele homozygotes (*r*=-0.05). This GEI effect was not present when the sample was stratified by sex (**Fig. 5B**). It arose in the combined analysis (males and females) as the QTL was sex-biased and had a larger additive effect on protein levels in males than in females (β=1.2 and 0.8 per-allele increase in transformed PAEP levels, respectively; both *p*<2.2×10^-16^). This difference is highlighted by the genotype-by-sex interaction analyses described above (*p*=4.7×10^-223^). Males with the T-allele therefore had higher protein levels than females as well as higher body weights. This forced a positive correlation between the protein and phenotype in the combined sample. The slight negative correlation in major allele homozygotes was due to males having slightly lower PAEP levels and higher body weights at this genotype (**Fig. 5B**).

**Fig. 5.**
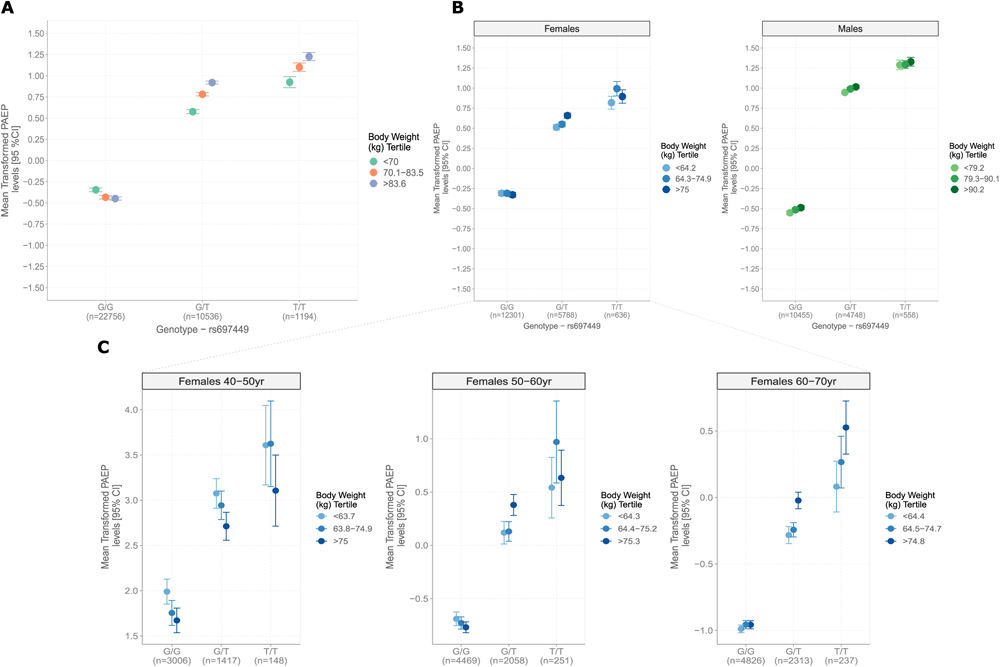
Relationships between glycodelin levels and body weight are influenced by age, sex and genotype. **(A)** Mean transformed glycodelin levels (PAEP, closed circles) with 95% confidence intervals (vertical bars) are shown according to tertiles of body weight (left-side, in kilograms, kg) and rs697449 genotype. **(B)** Relationship between glycodelin and body weight across genotypes as shown in (A) but stratified to females (blue) and males (green). **(C)** Relationship between glycodelin and body weight in females as shown in (B) but stratified into three separate decades of life.

Age also masked associations between PAEP levels and body weight in females (**Fig. 5C**). The age range of UKB-PPP participants spanned three separate decades of life at the study baseline (40-50, 50-60 and 60-70 years). Negative correlations between PAEP and body weight were observed across genotypes in the youngest age group (40-50 years, range of *r*=[-0.06,-0.13]). This age group also showed the highest glycodelin levels (**Fig. 5B**). Weak-to-moderate positive correlations were observed in the older age groups (range of *r*=[0,0.10] and [0.03,0.30]). The opposing associations across age groups nullified one another in the main analyses displayed in **Fig. 5B**. Interactions with genotype were also detected only in those aged 50-60 and 60-70 years (**Supplementary** Fig. 1). The age-dependent association between PAEP and body weight remained pronounced when the sample was further divided into 5-year age bins, with an apparent switch-point around 55 years of age (**Supplementary** Fig. 2). By contrast, associations between PAEP and body weight were consistent across age in males.

A three-term interaction between age, sex and genotype associated with 24 proteins at *p*<0.05, which included PAEP (*p*=4.8×10^-5^). These proteins associated with 30 metabolic, 13 physical and one lifestyle trait in GEI tests (**Supplementary Tables 11-13**). Age, sex and genotype may therefore together influence relationships between these plasma proteins and environmental exposures, as illustrated by PAEP and body composition.

### Gene-environment interactions explain why some vQTLs lack main effects on protein biomarkers

vQTLs may lack genetic main effects if the variant positively correlates with protein levels in one stratum of an exposure and negatively in other strata. The opposing effect sizes preclude a marginal main effect and produce a ‘directionally discordant’ GEI ^17^. A similar proportion of vQTLs with and without main effects at *p*<3.4×10^-11^ participated in conditional GEIs (15/65 or 23.1% and 137/618 or 22.2%, respectively). However, 13 of the 15 vQTLs (86.6%) without additive main effects were involved in directionally discordant interactions, compared to only 4/137 (2.9%) of those with additive main effects (**Supplementary Table 14**). Indeed, directionally discordant interactions likely explained why 13 of the wider 65 vQTL-only sites lacked additive main effects.

A clear example of this effect was observed at the *trans* indel in *FLT3* (rs147072313, fms-related tyrosine kinase 3), which associated with the variance of its ligand (FLT3LG). The binding of FLT3LG to cell-surface FLT3 promotes monocyte proliferation (**Fig. 6A**) ^21^. FLT3LG levels associated with a genotype-by-monocyte count interaction in this sample (*p*=6.5×10^-11^). rs147072313 positively correlated with FLT3LG levels within those assigned to the highest tertile of monocyte count and negatively in the lowest, likely precluding a main effect (**Fig. 6B**). The additive main effect was almost null (*p=*0.95, shown previously in **Fig. 2A**).

**Fig. 6.**
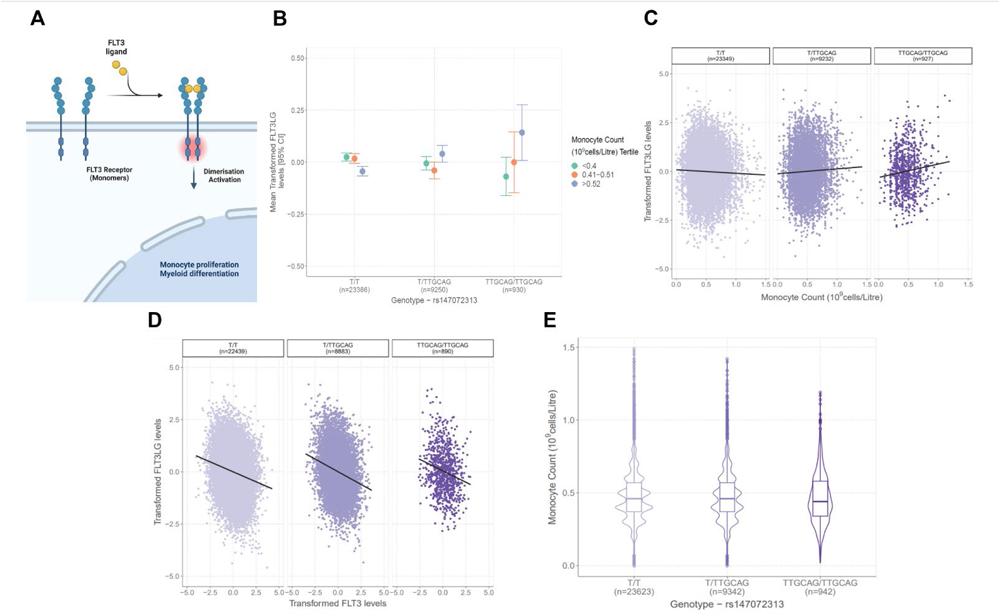
Variance QTL in FLT3 receptor gene modifies the relationship between FLT3 ligand levels and monocyte count. **(A)** Schematic diagram showing the role of FLT3 receptor and FLT3 ligand (FLT3LG) binding in monocyte proliferation. **(B)** Mean transformed FLT3LG levels (closed circles) with 95% confidence intervals (vertical bars) are displayed according to tertiles of monocyte count and rs147072313 genotype. Of note, the wide confidence intervals in those homozygous for the indel reflect the small sample size of this group. They represent confidence intervals for the mean of the protein levels across strata. They do not reflect the variance of the protein level, which is instead visualised in (C) and most clearly illustrated via boxplots in **Fig. 2A**. (C) displays full distributions of FLT3LG levels and their correlation with monocyte count. **(C)** The correlation between transformed FLT3LG levels and monocyte count differs according to genotype at rs147072313 giving rise to a gene-environment interaction. Here, it is also apparent that the vQTL confers reduced variance in FLT3LG in carriers of the indel. **(D)** The association between circulating FLT3LG levels and FLT3 receptor levels is not influenced by genotype at rs147072313. **(E)** Violin plots show that rs147072313 genotype does not have an additive main effect on monocyte count. QTL, quantitative trait locus. Image created using Biorender.com.

**Fig. 6C** shows that the indel was associated with decreased variance of transformed FLT3LG levels (*p*=2.1×10^-16^, also visualised previously in **Fig. 2A**). The GEI revealed a weak negative correlation between measured FLT3LG levels and monocyte count in major allele carriers (T-allele, *r*major=-0.03). A weak positive correlation was observed in heterozygotes (*r*het*=*0.03) and a stronger positive correlation was observed in indel homozygotes (*r*minor=0.11, **Fig. 6C**). This suggested that the effect of FLT3LG on monocyte count could be linked to rs147072313 genotype. However, the relationship between FLT3LG and FLT3 levels was consistent across genotypes, which suggested that ligand-receptor binding was preserved (**Fig. 6D**). Genotype also did not associate with mean differences in monocyte count (β=-0.001 per T-allele, *p*=0.59, **Fig 6E**). Therefore, the indel was not associated with an overall detrimental effect on monocyte count. The relationship was specific to monocytes when considering eight other blood cell types (**Supplementary** Fig. 3).

## Discussion

In this study, we utilised one of the world’s largest proteomic datasets and performed the first genome-wide vQTL association study on blood levels of approximately 1,450 proteins. We identified 1,400 GEIs across 142 blood proteins. We further highlighted GEIs that are either masked by or attributed to age and sex differences, and others that would not have been detected by conventional proteogenomic strategies.

Many of the exposures in this study do not represent behavioural or environmental exposures external to an organism. However, they were included due to prior evidence for their relevance to GEIs that involve blood-based biomarkers as well as interactions between the proteome and metabolome ^17,22^. For instance, we observed positive correlations between LDLR and LDL cholesterol in carriers of the major C-allele at rs75627662, which is located near *APOE*. The association was attenuated in individuals with two copies of the minor T-allele. Further, the T-allele was associated with elevated LDLR levels in our study, and has been linked to adverse lipid profiles and reduced expression in whole blood of genes within the *TOMM40/APOE/APOC1* locus ^23,24^. APOE is an important component of several lipoprotein particles including LDLR. Typically, hepatic cell-surface LDLR clears cholesterol from the circulation. Therefore, a negative association between membrane-bound LDLR and cholesterol may be anticipated ^25^. By contrast, positive correlations between soluble or circulating LDLR and LDL cholesterol have been reported previously ^26–29^. One possible mechanism that explains the positive association entails shedding of cell-surface LDLR. Shedding increases the soluble form of the receptor and raises cholesterol levels due to the reduced availability of cell-surface proteins to clear circulating particles. Our findings may suggest that normal LDLR post-translational regulatory mechanisms are compromised in T-allele carries. Further, this relationship may be linked to genetic variation within the *TOMM40/APOE/APOC1* cluster. Elevated LDLR levels in minor allele carriers could also likely underscore dysregulated receptor turnover and lipid metabolism. However, it is important to note that measured levels do not necessarily inform about activity and these findings warrant further investigation in follow-up mechanistic *in vitro* and *in vivo* studies.

Associations involving glycodelin and physical traits were attributed to sexual dimorphism in body composition and sex-biased QTL effects. By contrast, associations between glycodelin and body composition were masked by age, but only within females. Females under 55 years of age showed negative correlations between glycodelin and body mass whereas those over 55 years exhibited positive correlations. Glycodelin has four major glycoforms (-A,-S,-F and –C) with prominent roles in reproduction, pregnancy and immune function ^30^. Plasma glycodelin levels are highest in females between 40-45 years of age with a steady reduction until 55 years, after which plasma profiles resemble those of male counterparts ^4^. Over 90% of females in UKB self-reported menopausal onset by 55 years (data field 3581). Of note, genetic variation within *PAEP* is not linked to differences in body composition. However, our data suggest that the role of glycodelin within the homeostatic landscape may become altered following the onset of menopause. This provides one example whereby age, sex and genotype interact to modify the association between biomarkers and health-related phenotypes. These findings are important as previous GEI studies often only correct the biomarker for demographic variables and include sexually dimorphic exposures in association tests. They also do not often perform stratified analyses and therefore may be agnostic to age variation and sex. This finding further underscores the need for large samples to enable stratified analyses that can inform biomarker research in understudied groups.

In addition to demographic variables, we identified exposures that explained why some loci affect the variance of protein biomarkers only. For instance, the effect of a *trans* indel within *FLT3* on the distribution of its ligand FLT3LG was explained by an interaction with monocyte count. A conventional model suggests that lower circulating FLT3LG levels serve as a proxy for higher receptor-bound ligand. In turn, higher receptor-coupled ligand produces higher monocyte count, leading to a negative correlation between free ligand and cell count ^31^. An unexpected positive correlation was observed in indel carriers. However, monocyte count and receptor-ligand binding were stable across genotype groups. This suggested that the indel may instead impact downstream ligand-mediated signalling processes without exerting overall detrimental effects on blood cell profiles. GEIs are one possible explanation for a vQTL that lacks a main effect, which may instead reflect phantom vQTLs ^32,33^, phenotypic dispersion induced by selection and epistasis^33^. In particular, 16 vQTLs that lacked main effects were located within the pleiotropic *HLA* and *ABO* regions, which could underscore gene-gene interactions. Complex interactions between multiple environmental exposures or inaccurately recorded phenotypes could also have precluded GEI detection, in particular within the large correlated constellation of phenotypes studied.

Large biobank efforts have successfully detected genetic underpinnings of the human proteome, defining variants that associate with mean differences in protein levels. Whilst variants with additive main effects (i.e. non-vQTL sites) can be used in GEI tests, we observed that the proportion of vQTLs participating in a GEI was five-fold higher than corresponding main effect sites. Therefore, this study provides additional evidence that vQTLs increase the likelihood of identifying an underlying gene-environment interaction when compared to main effect loci. The higher number of main effect loci over vQTLs reflects the greater statistical power of standard regression methods when compared to Levene’s test. By contrast, the power of GEI tests may increase when using vQTLs rather than main effect loci. Variance QTLs in particular may be susceptible to differences in phenotype transformations owing to their reliance on the distribution of the trait. This could result in some vQTLs representing artefacts of the nominated transformation ^34,35^. However, our sensitivity analyses suggested that the majority of associations observed in our study were robust to phenotype pre-processing and transformation.

The two-stage design and stepwise conditional analyses have been applied previously ^15–17,36^. However, our study advances beyond the existing literature by (i) conducting the first vQTL analyses on over 1,400 blood proteins, (ii) considering the largest number of biomarkers in vQTL-GEI studies to date and crucially, (iii) demonstrating how demographic variables (e.g. age and sex differences) may mask or induce observed GEIs. The latter addition provides novel biological insights alongside crucial methodological considerations for GEI discovery as combined multi-omic and phenomic resources expand. Importantly, others will be able to utilise our prioritised set of vQTLs and extract their genotypes to perform GEI tests with phenotypes of interest in their study.

This study has a number of limitations. First, an external replication set was not available as the set of proteins and phenotypes are presently unique to UKB. Second, the majority of participants were of European ancestry. It is not possible to clearly generalise gene-environment effects to other ancestry groups given potential differences in both genetic and environmental profiles. Third, specific quality control parameters could not feasibly be applied to all phenotypes tested. Additional confounders and phenotype-specific transformations may need to be considered in follow-up or mechanistic studies.

## Conclusion

The study complements existing proteogenomic efforts by considering additional distributional properties of the proteome whilst cataloguing biologically informative examples of its interaction with the genome, metabolome and phenome. Our datasets of variance QTL effects and GEIs establish unmet resources in the pursuit of accelerating biomarker discovery and validation.

## Online Methods

### UK Biobank Study

UKB is a prospective, population-based cohort of approximately 500,000 individuals aged between 40-69 years at recruitment ^37^. Recruitment took place between 2006 and 2010. Here, a subset of the UKB sample was utilised, which was defined by The UKB Pharma Proteomics Project or UKB-PPP consortium. The consortium comprises 13 biopharmaceutical companies, which funded the generation of blood-based proteomic data. The UKB-PPP sample includes 54,306 participants and consists of (i) a randomised subset of 46,673 UKB participants at the baseline visit, (ii) 6,385 individuals at the baseline selected by the UKB-PPP consortium members and (iii) 1,268 individuals who participated in the COVID-19 repeat imaging study.

### Protein measurement in UK Biobank

Blood samples from 54,306 UKB-PPP participants were analysed using the Olink Explore 1536 platform. The platform uses Proximity Extension Assay ^38^ and measured 1,472 protein analytes across four Olink panels (Cardiometabolic, Inflammation, Neurology and Oncology). The analytes reflect 1,463 unique proteins. EDTA-treated plasma samples (60µl) were serially diluted to 1:10, 1:100 and 1:1000 and transferred to 384-well plates. Samples were processed in eight batches (termed batches 0-7) and incubated with antibodies overnight at -4°C. Olink’s inbuilt quality control (QC) workflow returned Normalized Protein eXpression (NPX) values, which is a relative quantification unit on a log-2 scale. Full details on protein measurement and QC are available in **Supplementary Methods**. Protein data were available for 54,189 individuals following QC.

### Genotyping in UK Biobank

The UKB genotype dataset includes 488,377 participants. Of these, 49,950 individuals were genotyped on the Applied Biosystems UK BiLEVE Axiom™ Array and 438,427 participants were genotyped on the closely related UK Biobank Axiom™ Array ^37^. Here, we followed the genotype QC process of Sun *et al.* (2022) in order to enable direct comparisons to a conventional main effect QTL analysis using the same sample ^4^. Briefly, UKB genotype data were imputed to the Haplotype Reference Consortium ^39^ and UK10K ^40^ reference panels. Imputed genetic variants were filtered for INFO>0.7 and minor allele count>50, and chromosome positions were lifted to hg38 build using LiftOver ^41^. Variants with a genotyping rate >99%, Hardy-Weinberg equilibrium test *p*>10^−15^ and <10% missingness were retained. Sun *et al.* (2022) utilised variants with minor allele frequency (MAF)>1%. We applied a higher threshold of MAF>5% in accordance with the workflow of Wang *et al.* for vQTL analyses ^15^. Following QC, 6,815,574 variants remained. There were 53,752 individuals with paired genotype and protein data.

The UKB-PPP sample was separated into discovery (n=35,571) and replication subsets (n=18,181) as per the design of Sun *et al.* (2022) ^4^. The discovery set included participants who were of European ancestry and present in Olink measurement batches 1-6. The remaining samples comprised the replication set and included 14,706 White, 1,225 Black/Black British, 998 Asian/Asian British, 148 Chinese, 339 Mixed, 613 Other and 152 individuals with missing self-reported ethnic backgrounds (as defined by data field 21000).

### Variance QTL association studies

vQTL association studies were performed using the vQTL suite in OSCA ^42^. Levene’s test with median was applied. The false-positive rate of this test has been shown to be well-calibrated across simulated data in comparison to other commonly-used vQTL methods ^15^.

In the discovery cohort, rank-based inverse normal transformed protein values (NPX) were regressed onto age, age^2^, sex, age*sex, age^2^*sex, batch, UKB study centre, UKB genotype array, time between blood sampling and measurement and 20 genetic principal components. One additional covariate was included in the replication set, which indicated whether samples were pre-selected by consortium members or as part of the COVID imaging study. Summary data for covariates are available in **Supplementary Table 15** and their associations with protein levels are shown in **Supplementary Tables 16 and 17** for discovery and replication sets, respectively. The preparation of protein data was aligned as closely as possible to the corresponding main effect QTL study by Sun *et al.* ^4^. Of note, Sun *et al.* did not adjust protein levels prior to genetic association studies and instead included fixed-effect covariates. This was not possible in our study as the Levene’s test module did not permit fixed-effect covariates. Residuals were standardised to Z-scores and entered as dependent variables. Additively-coded genotype status was included as the independent variable.

A Bonferroni-corrected significance threshold of *p*<3.4×10^-11^ was applied (*p*<5×10^-8^ adjusted for 1,463 proteins). Primary associations were defined by clumping variants ±1Mb around significant vQTLs using PLINK ^43^ with the exception of the HLA locus (chromosome 6: 25.5-34.0 Mb). The larger HLA locus was considered as one region due to its complex linkage disequilibrium structure.

### Preparation of phenotypes for gene-environment interaction tests

Prior to QC, 601 candidate phenotypes were considered for GEI tests. Phenotypes with excessive missingness (i.e. >20%, n=472) and insufficient heterogeneity (n=1) were removed in line with the protocol of the PHESANT algorithm ^44^. An additional fourteen phenotypes were removed due to lack of relevance to health outcome testing (i.e. were technical or QC variables). There were 114 phenotypes following QC. Categorical variables were converted into binary or ordinal phenotypes as appropriate. Continuous variables were rank-based inverse normal transformed. The phenotypes or ‘exposures’ consisted of 63 metabolic, 46 physical, four lifestyle and one technical variable (season of blood draw). Further information on phenotype selection and their preparations are available in **Supplementary Methods.** Principal component analysis was performed using the R function *prcomp* on all 114 phenotypes in the discovery set, which revealed 13.5 effective phenotypes for multiple testing correction.

### Gene-environment interaction tests

Linear regression models tested whether the levels of a given protein were associated with an interaction between its vQTL (stage one) and each of 114 possible exposures (stage two):

> *Protein levels (standard dised residuals) ∼ SNP (0,1,2) ∗ exposure + SNP + exposure*

No fixed-effect covariates such as age and sex were included in GEI association models as protein levels were already corrected for relevant covariates in stage one, and in keeping with prior GEI studies ^17^. A Bonferroni-corrected threshold of *p*<5.4×10^-6^ was applied (*p*<0.05 adjusted for 683 vQTLs and 13.5 effective exposures). By contrast, vQTL association tests in stage one were adjusted for the total number of proteins rather than their effective number in order to match the threshold used in Sun *et al.* and to ensure appropriate cross-study comparisons. As described in Westerman *et al.* ^17^, conditional GEI tests were performed in stage two in order to further account for the correlation structure between related phenotypes. All GEIs for a given protein that withstood multiple testing correction were brought forward to stepwise conditional analyses. Interaction effects for a given protein were re-tested while iteratively conditioning on the interaction that involved the most significant exposure (i.e. smallest *p*-value for a given protein). The process was repeated until no further GEIs could be considered. Interactions with age and sex were also investigated in addition to 114 candidate exposures. In standard GEI tests (i.e. with the 114 candidate exposures), protein levels were corrected for age, age^2^ or sex and interaction terms involving these variables as described in the preceding sections. However, protein levels were adjusted only for technical factors (e.g. batch and study centre) and genetic PCs when age and sex were included as possible exposures.

## Data availability

Genome-wide vQTL summary statistics and GEI association data will be made available via an open-access repository upon publication. Proteomics data are available in UK Biobank under return dataset [return dataset ID and URL will be provided upon publication].

## Code availability

All code is available with open access at the following GitHub repository: https://github.com/robertfhillary/vqtls-uk-biobank.

## Acknowledgements

This research was funded in whole, or in part, by the Wellcome Trust [108890/Z/15/Z]. For the purpose of open access, the author has applied a CC BY public copyright licence to any Author Accepted Manuscript version arising from this submission.

We thank the participants, contributors, and researchers of UK Biobank for making data available for this study – with special thanks to Lauren Carson, John Busby, Naomi Allen and Rory Collins for making the study possible. We are grateful to the research & development leadership teams at the thirteen participating UKB-PPP member companies (Alnylam Pharmaceuticals, Amgen, AstraZeneca, Biogen, Bristol-Myers Squibb, Calico, Genentech, Glaxo Smith Klein, Janssen Pharmaceuticals, Novo Nordisk, Pfizer, Regeneron, and Takeda) for funding the study. We thank the Legal and Business Development teams at each company for overseeing the contracting of this complex, precompetitive collaboration – with particular thanks to Erica Olson of Amgen, Andrew Walsh of GSK and Fiona Middleton of AstraZeneca. Finally, we thank the team at Olink Proteomics (Philippa Pettingell, Klev Diamanti, Cindy Lawley, Linda Jung, Sara Ghalib, Ida Grundberg and Jon Heimer) for their consistent logistic support throughout the project – with special thanks to Evan Mills for co-championing the project and leading internal activities at Olink.

R.E.M. is supported by Alzheimer’s Research UK major project grant **ARUK-PG2017B−10** and Alzheimer’s Society major project grant **AS-PG-19b-010**. D.A.G. is supported by the Wellcome Trust Translational Neuroscience programme [**108890/Z/15/Z**]. R.F.H is supported by a British Heart Foundation Immediate Fellowship [**FS/IPBSRF/22/27042**].

## Author contributions

R.F.H., D.A.G., Z.K., T.M., H.R., R.E.M., C.N.F. and B.B.S. conceptualised the study design and consulted on methods and results. R.F.H performed all main analyses. T.L., K.F. and H.M. performed quality control or prepared the proteomics dataset. All authors reviewed and approved of the manuscript.

## Ethics declarations

All participants provided informed consent. This research has been conducted using the UK Biobank Resource under approved application numbers 65851, 20361, 26041, 44257, 53639, 69804.

## Materials and correspondence

All correspondence and material requests should be sent to Dr Benjamin Sun at. For any further correspondence requests please contact Prof Riccardo Marioni at.

## Supplementary information captions

- **Supplementary Tables - Table S1**. Genomic inflation factors from all 1,472 genome-wide variance QTL association studies. **Table S2**. Independent variance QTLs at *p*<3.4×10^-11^ (LD-pruned, Bonferroni-corrected threshold). **Table S3**. Variance QTL association statistics in replication set. **Table S4**. Look-up analyses for main effect QTLs in Sun *et al.* (2022). **Table S5**. Summary data for phenotypes used in gene-environment interaction or GEI tests. **Table S6**. All significant GEI associations with variance QTLs at *p*<5.4×10^-6^ (Bonferroni-corrected threshold). **Table S7**. All significant GEI associations with main effect QTLs at *p*<5.4×10^-6^ (Bonferroni-corrected threshold). **Table S8**. Conditionally significant GEI associations in discovery set at *p*<5.4×10^-6^ (Bonferroni-corrected threshold). **Table S9**. Conditionally significant GEI associations in replication set. **Table S10**. Associations between protein levels and age-by-genotype and sex-by-genotype interaction terms. **Table S11**. Associations between protein levels and a three term age-by-sex-by-genotype interaction. **Table S12**. GEI associations stratified by sex and age (10-year bins) for proteins from Table S11. **Table S13**. GEI associations stratified by sex and age (5-year bins) for proteins from Table S11. **Table S14**. Direction of associations between genotype and protein levels across different strata of exposures from Table S8. **Table S15**. Summary data for demographic variables and covariates in UKB-PPP sample. **Table S16**. Associations between proteins and covariates in the discovery set. **Table S17**. Associations between proteins and covariates in the replication set. **Table S18**. Sensitivity variance QTL analyses using phenotype preparation method recommended by Wang *et al.* (2019). **Table S19**. Sensitivity variance QTL analyses using protein levels that were regressed on partner phenotype(s) in conditional GEI associations. Tables S18 and S19 are referred to in sensitivity analyses that are described in full in **Supplementary Note**.
- **Supplementary Note -** Details sensitivity analyses for protein data preparation and quality control.
- **Supplementary Figures** - **Fig. S1**. Gene-environment interactions between glycodelin and body composition stratified by sex and ten-year age bins. **Fig. S2**. Relationship between glycodelin and body composition stratified into five-year age bins within males and females separately. **Fig. S3**. Correlation between FLT3LG levels and eight different blood cell types stratified by genotype at a variance QTL for FLT3LG levels.
- **Supplementary Methods** - Further details for (i) **sample selection**, (ii) **proteomic profiling and quality control**, (iii) **vQTL association studies** and (iv) **phenotype selection for gene-environment interaction association tests**.

