## Supplementary Note for "Systematic discovery of gene-environment interactions underlying the human plasma proteome in UK Biobank"

**Supplementary Note**. The file pertains to a Supplementary Note on sensitivity analyses for protein data preparation and quality control and is associated with the manuscript ‘*Systematic discovery of gene-environment interactions underlying the human plasma proteome in the UK Biobank*’ by Hillary *et al*.

**Variance QTLs and gene-environment interactions withstand sensitivity analyses**

The false-positive rate of Levene’s test may be inflated in the presence of QTL effects if the phenotype was non-linearly transformed (e.g. RINT). As sensitivity analyses, we re-estimated vQTL associations when preparing phenotypes in line with guidelines by Wang *et al.* to avoid these potential biases ^1^ . Here, protein levels (NPX values) were regressed onto age, age^2^, batch, UKB study centre, UKB genotype array, time between blood sampling and measurement and 20 genetic principal components within males and females separately. Residuals were then combined and standardised to Z-scores. Relative effect sizes were correlated 90.4% (95% CI=[88.9%, 91.9%]) between tests that used phenotypes prepared with RINT (i.e. main analytical strategy) and those prepared according to Wang *et al.* (sensitivity analysis). Of the 683 primary vQTLs identified in this study, 639 (93.6%) remained significant at *p*<0.05 in this sensitivity analysis with 551 (80.7%) remaining at *p*<3.4x10^-11^ (**Supplementary Table 18**).

Furthermore, as described in Zhang *et al.* (2016) ^2^ and Westerman *et al.* (2022) ^3^, the two-stage strategy employed in this study may have inflated type I error if an exposure associates with protein levels. In this case, test statistics from stage one and two might be correlated. Therefore, we repeated vQTL analyses in stage one but this time each protein that participated in a conditionally significant GEI in stage two was regressed onto the exposure(s) it associated with in stage two. Association statistics were then re-estimated for the 152 vQTLs that were implicated in 215 conditional GEIs within stage two. Relative effect sizes were correlated 98.8% (95% CI=[98.3%, 99.2%]) with those in the main analytical strategy. Further, 148 vQTLs (97.4%) remained significant at *p*<0.05 in this exposure-adjusted sensitivity analyses and 132 associations (86.8%) remained at *p*<3.4x10^-11^ (**Supplementary Table 19**).
