## Supplementary Figures for "Systematic discovery of gene-environment interactions underlying the human plasma proteome in UK Biobank"

**Supplementary Figures.** The file pertains to Supplementary Figures for the manuscript ‘*Systematic discovery of gene-environment interactions underlying the human plasma proteome in UK Biobank*’ by Hillary *et al.*

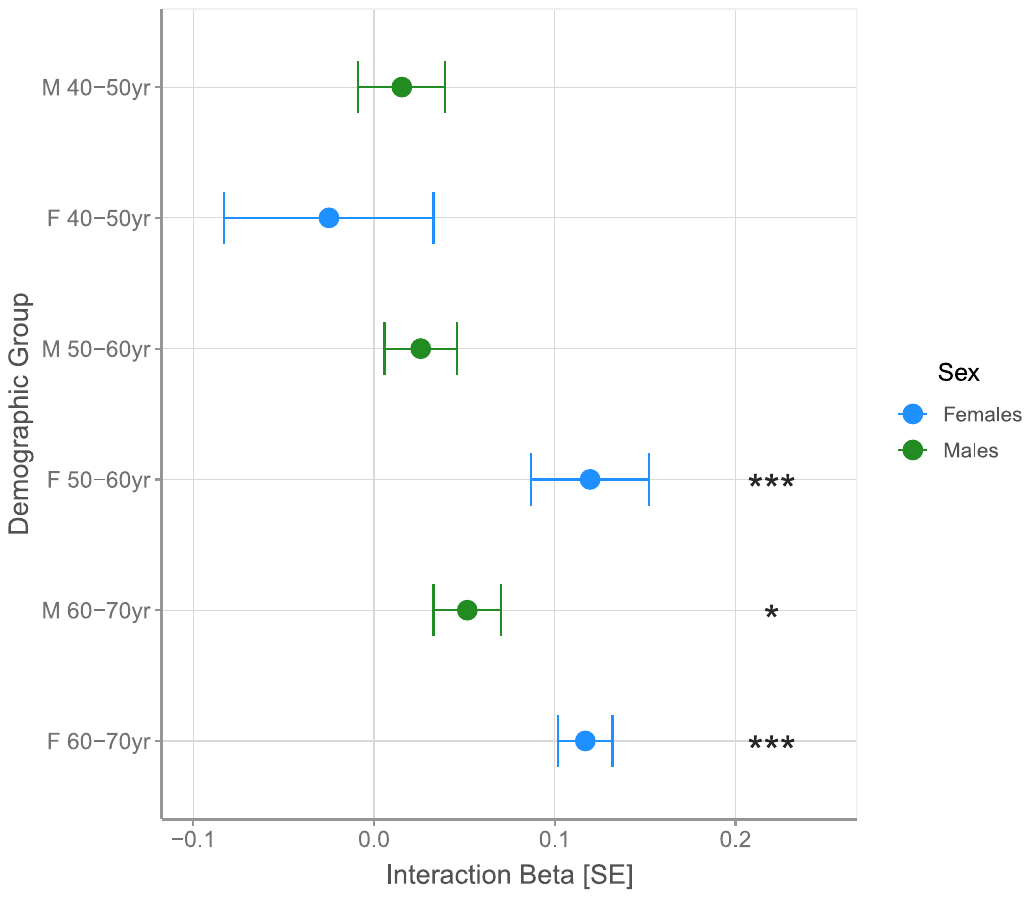

**Fig. S1**. **Gene-environment interactions between glycodelin and body composition stratified by sex and ten-year age bins**. Effect sizes (closed circles) and standard errors (vertical bars) for genotype-by-body weight interaction effects on PAEP levels, stratified by age and sex in UK Biobank. F, females; M, males; SE, standard error.

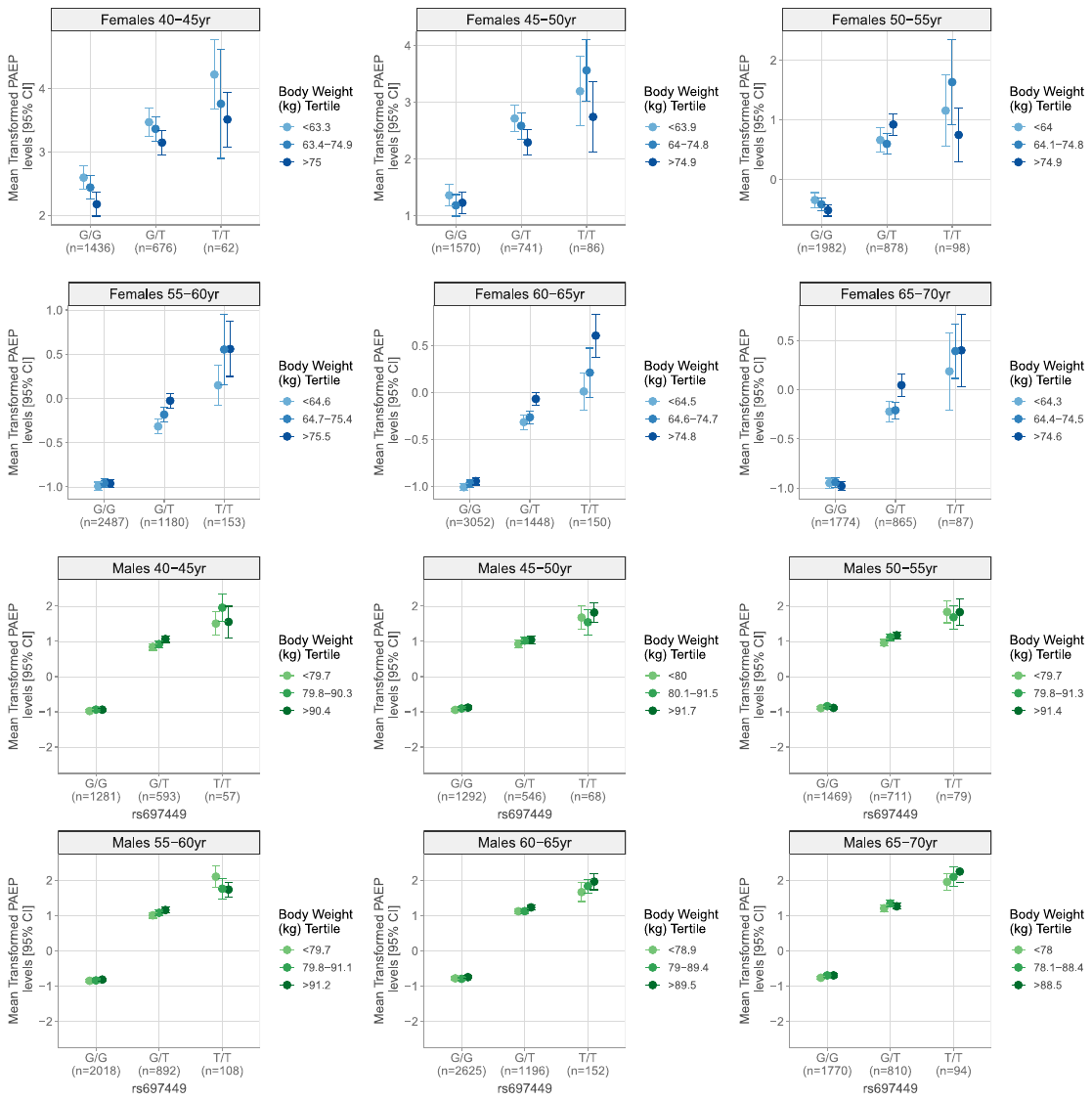

**Fig. S2**. **Relationship between glycodelin and body composition stratified into five-year age bins within males and females separately**. Mean transformed glycodelin levels (PAEP, closed circles) with 95% confidence intervals (vertical bars) are shown according to tertiles of body weight (in kilograms, kg) and rs697449 genotype. The plots are stratified into females (blue) and males (green) with further subdivisions by five-year age bins. CI, confidence interval.

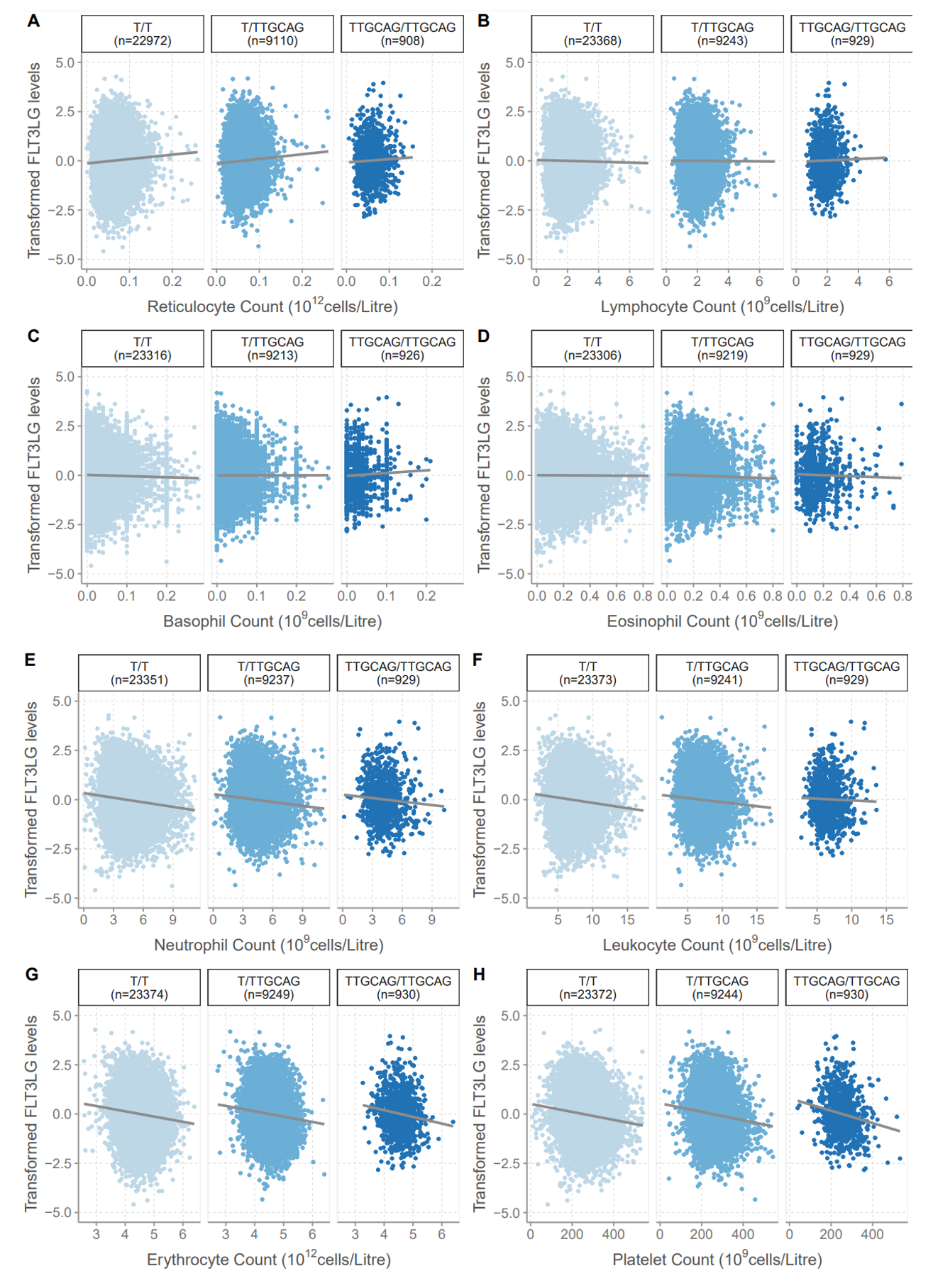

**Fig. S3. Correlation between FLT3LG levels and eight different blood cell types stratified by genotype at a variance QTL for FLT3LG levels**. **(A)** Reticulocyte counts. **(B)** Lymphocyte counts. **(C)** Basophil counts. **(D)** Eosinophil counts. **(E)** Neutrophil counts. **(F)** Leukocyte counts. **(G)** Erythrocyte counts. **(H)** Platelet counts. QTL, quantitative trait locus.
