## Supplementary Methods for "Systematic discovery of gene-environment interactions underlying the human plasma proteome in UK Biobank"

**Supplementary Methods**. The file pertains to Supplementary Methods for the manuscript ‘*Systematic discovery of gene-environment interactions underlying the human plasma proteome in UK Biobank*’ by Hillary *et al*.

**Sample selection**

In the first wave of selection, 5,500 samples collected from participants at the study baseline were pre-selected by The UK Biobank Pharma Proteomics Project (UKB-PPP) consortium members. Following this, 44,502 additional and representative samples were selected from baseline visits via a stratified selection on age, sex and UKB study centre. This strategy was employed to reduce the number of plates for the picking process. There were 50,002 samples following the first wave of selection. In the second wave of selection, 7,000 samples were selected. This included 1,020 samples selected by the consortium and 3,637 participants in the UKB COVID-19 repeat imaging study. Of these 3,637 participants, 1,270 samples from baseline, pre-COVID and post-COVID imaging visits were included where possible. An additional 2,343 baseline samples were included in the second stage and were selected randomly as in the first stage in order to optimise the inclusion of selection locations required for the COVID imaging and consortium selected samples. Consortium members chose samples that were enriched for specific diseases of interest. Day of week for sample collection, deprivation index and participant ethnicity were representative of the wider UK Biobank cohort. Full inclusion and eligibility criteria are detailed in: <https://biobank.ndph.ox.ac.uk/showcase/showcase/docs/casecontrol_covidimaging.pdf>.

**Proteomic profiling and quality control**

In the Proximity Extension Assay, blood samples are incubated in the presence of proximity antibody pairs linked to DNA reporter molecules. Upon binding of an antibody pair to their corresponding antigen, the respective DNA tails form an amplicon by proximity extension, which can be quantified by high-throughput real-time PCR. This method limits the reporting of cross-reactive events. Amplicons were combined across each of four distinct abundance groups resulting in one well of amplicons per sample. Four unique 96-index plates were added to every four sample plates followed by a second PCR reaction, which enabled all samples in a plate to be combined into one library per panel. Each library was bead-purified using a Bioanalyzer. Four sample plates from four identical panels were pooled, denatured and sequenced on individual lanes on the Novaseq600 using S4 flow cells v1.5 (35 cycles) and 384-samples and 384-assays measured. Counts of known sequences were translated into Normalized Protein eXpression (NPX) values within Olink’s MyData Cloud Software.

Raw data were generated for 54,309 individuals. Olink’s inbuilt QC system includes three controls (incubation, extension and amplification controls) that are spiked into every sample and to each abundance block. Each sample also includes external controls in the twelfth column and a triplicate negative control is added to aid in calculating limits of detection. The proteins IL6, IL8 (CXCL8), and TNF were present across several panels and helped to assess consistency and correlations of protein measurements. Olink samples were divided into eight batches. Samples were randomised across 96-well plates. The first seven batches (labelled batch 0-6) included participants from UKB baseline. The eighth batch (labelled batch 7) contained samples from the baseline and the COVID imaging study. Twenty-five plates from batch 7 included baseline samples only, whereas 94 plates contained both baseline and COVID imaging visit samples. Within each plate, there were 87 biological samples, 2 QC controls, 3 negative controls to compute the baseline assay level of each plate and 3 Olink plate control samples that were used for the normalisation of protein expression. NPX values were calculated by subtracting the assay-specific median value of plate controls from the log_2_ ratio of counts of each assay-sample pair to the counts of the extension control. This was performed separately for baseline and COVID imaging samples. This provided plate normalized NPX values for both sets. For baseline samples, batch-specific median NPX values were added per assay in order to account for batch-to-batch variation. Data were then normalised within each of the batches. Adjustment factors were calculated using batch 1 as the reference batch and added to baseline-only batches. Adjustment factors for batch 7 were computed from the assay-specific median of the pairwise differences between bridge samples in batches 0-6 and batch 7. These adjustment factors were added to NPX values within batch 7.

NPX values were available for 58,699 samples and 54,309 individuals. Removing control, unprocessed and withdrawn samples left 58,362 samples and 54,306 individuals. Samples were also removed if they were (i) Olink QC failures, (ii) outliers that had a standardised principal component (PC) 1 or 2 beyond 5 standard deviations from the mean (zero-standardised PCA), (iii) outliers with a median NPX greater than 5 standard deviations from the mean or an interquartile range (IQR) greater than 5 standard deviations from the mean IQR and (iv) flagged by QC or assay warnings. These criteria left measurements for 54,189 individuals, comprising 58,249 samples (due to some participants providing repeat samples). Baseline samples for these 54,189 individuals were taken forward for use in the present study.

**vQTL association studies**

vQTL effect sizes and their standard errors are not natively produced by Levene’s test. First, a direction of effect is inferred by regressing absolute deviations of phenotype values from the median onto additively-coded genotype values (0, 1 or 2). Second, effect sizes and standard errors are back-transformed using the sign of the effect’s direction and the test’s *p*-value.

**Phenotype selection for gene-environment interaction association tests**

The selected traits were included given prior evidence of GEIs with anthropometric and adiposity traits ^1-3^, and interactions between the proteome and metabolome ^4,5^. We also included four common lifestyle risk factors: alcohol intake frequency (6-category ordinal variable), education (binarised to having completed college/university education), the Townsend deprivation index (higher scores indicating greater deprivation) and smoking status (3-category ordinal variable). Lastly, season of blood draw was included as a sole technical exposure due to its potential influence on inter-individual variation in protein measurements ^6^. Season of blood draw was based on the blood collection date and time (data-field: 3166) and binarised to ‘Summer/Autumn’ (June to November) and ‘Winter/Spring’ (December to May).
